# Impact of Moderate to Severe Mitral and Tricuspid Valves Regurgitation after Transcatheter Aortic Valve Implantation: A Multicenter Retrospective Study

**DOI:** 10.1101/2024.02.15.24302908

**Authors:** Bishoy Abraham, Michael Botros, Ayman Fath, Sara Kaldas, Michael Megaly, Chieh-Ju Chao, Reza Arsanjani, Chadi Ayoub, F. David Fortuin, John Sweeney, Patricia Pellikka, Vuyisile Nkomo, Mohamad Alkhouli, David Holmes, Said Alsidawi

## Abstract

Tricuspid regurgitation (TR) and mitral regurgitation (MR) are common valvular conditions encountered in patients undergoing transcatheter aortic valve implantation (TAVI). This retrospective study investigates the impact of moderate or severe TR and MR on all-cause mortality in one-year post-TAVI patients. Consecutive patients who underwent TAVI at our institution between 2012 and 2018 were identified. Patients were stratified into 2 groups based on valvular regurgitation severity: moderate/severe MR vs. no/mild MR, and moderate/severe TR vs. no/mild TR. Primary outcome was all-cause mortality at 1-year follow up, and secondary outcome was in-hospital death. Logistic regression analysis was conducted to assess the relationship between moderate/severe MR or TR and all-cause mortality at 1-year follow-up. We included a total of 1,071 patients who underwent TAVI with mean age 80.9 ± 8.6 years, 97% white, and 58.3% males. Moderate or severe MR group included 52 (4.88%) patients while mild or no MR group included 1,015 (95.12%) patients. There was no significant difference between both groups in TAVI procedure success rate (100% vs. 97.83%, p=0.283), in-hospital mortality (0 vs. 1.08%, p=0.450), or mortality at 1-year follow up (15.38% vs. 14.09%, p=0.794). In the logistic regression analysis, moderate or severe MR did not show significant correlation with all-cause mortality at 1-year follow up (OR 0.88, 95% CI [0.38 - 1.99], p=0.761). Moderate or severe TR group included 86 (8.03%) patients while mild or no TR group included 985 (91.97%) patients. There was no difference between both groups in TAVI procedure success (98.8% vs. 97.9%, p=0.54) or in-hospital mortality (0% vs. 1.1%, p=0.33). At 1-year follow up, patients with moderate or severe TR had higher mortality (26.7% vs. 13.2%, p=0.001) compared to patients with mild or no TR. In the logistic regression analysis, moderate or severe TR was correlated with higher all-cause mortality at 1-year (OR 1.89, 95% CI [1.07, 3.34], P=0.027). At 1 year follow up, moderate/severe TR, but not MR, is associated with higher mortality in patients underwent TAVI. Careful assessment of multi-valvular heart disease prior to the procedure is warranted.

## Introduction

The most common valve heart disease in the western world is degenerative aortic valve stenosis (AS) (1). A significant proportion of patients receiving transcatheter aortic valve implantation (TAVI) have moderate or severe mitral regurgitation (MR) and/or tricuspid regurgitation (TR). The combination of AS and significant other valvular regurgitation may impact the diagnosis, prognosis, and treatment (2).

An improvement in MR severity after aortic valve (AV) surgery is linked to improved outcomes and decreased mortality (2,3). However, a challenge arises due to the asynchronous progression of AS and MR, affecting the exact time of necessary intervention. Also, studies have revealed that up to 15% of patients diagnosed with severe AS also exhibit moderate or severe TR, underscoring the prevalence and clinical complexity of these concurrent conditions (4,5). Addressing both conditions requires a thorough assessment of valve anatomy and function, and understanding the underlying pathophysiological pathways to establish appropriate treatment methods and planning procedures (6,7,8).

Traditional open-heart surgery manages both AS and significant valve regurgitation simultaneously, reducing the risks associated with multiple interventions. In contrast, recent transcatheter techniques present a more flexible approach, enabling customized intervention time, and offering less invasive options for staged or combined procedures, especially for elderly patients, avoiding the need for traditional surgery. The guidelines advise against surgery for non-severe non-ischemic MR without structural issues, as treating aortic stenosis might occasionally relieve the MR. However, a lack of specific guidance for more significant MR and TR remains a topic of ongoing discussion, lacking clear guideline-based recommendations. (6,9,10,11,12). The independent impact of concurrent MR and TR on clinical outcomes following TAVI is still being researched and debated. This study aimed to investigate the impact of concomitant significant valvular disease on patients undergoing TAVI.

## Methods

We performed a retrospective observational study utilizing the Mayo Clinic National Cardiovascular Diseases Registry database, which included patients from three major academic medical centers in Rochester, MN, Phoenix, AZ, and Jacksonville, FL. We included all consecutive patients who underwent TAVI for severe aortic valve stenosis from January 2012 to December 2018. We used the last available pre-procedure transthoracic echo to identify other concomitant valvular abnormalities. Moderate or severe MR was defined as regurgitation volume >45 ml/beat, regurgitation fraction >40%, and/or effective regurgitation orifice area >0.30 cm^2^. Moderate or severe TR was defined as regurgitation jet area >5 cm^2^, vena contracta width >0.4 cm, and hepatic vein systolic flow blunting or reversal. We retrospectively collected data on the baseline characteristics of the included patients, echocardiographic parameters prior to the TAVI procedure, and the outcomes of interest. TAVI procedure success was defined as proper placement of the prosthesis with mean gradient <20 mmHg or peak velocity <3 m/s, and absence of procedural mortality. The primary outcome was all-cause mortality at 1-year follow up, and the secondary outcome was in-hospital death. We used STATA software version 17 for the statistical analysis. We reported categorical variables as numbers and percentages, and the continuous variables as mean ± standard deviation (SD). Categorical variables were analyzed using Pearson’s Chi-square or Fisher’s exact tests. Continuous variables were analyzed using two-sample t test or Mann-Whitney U test. P values were two-sided and P < 0.05 was considered. Multivariate analysis was conducted to identify variables associated with all-cause mortality at 1-year follow up. We included all known variables that may affect mortality such as age, sex, diabetes, hypertension, and prior MI. The Mayo Clinic institutional review board approved the study with patients’ consent waiver.

## Result

### Comparing moderate/severe MR vs. mild/no MR

We included a total of 1,067 patients who underwent TAVI, 52 patients (4.88%) had moderate/severe MR, and 1,015 patients (95.12%) had mild/no MR. **Table 1** summarizes the demographics and baseline patients characteristic of the two groups.

**Table 1.**
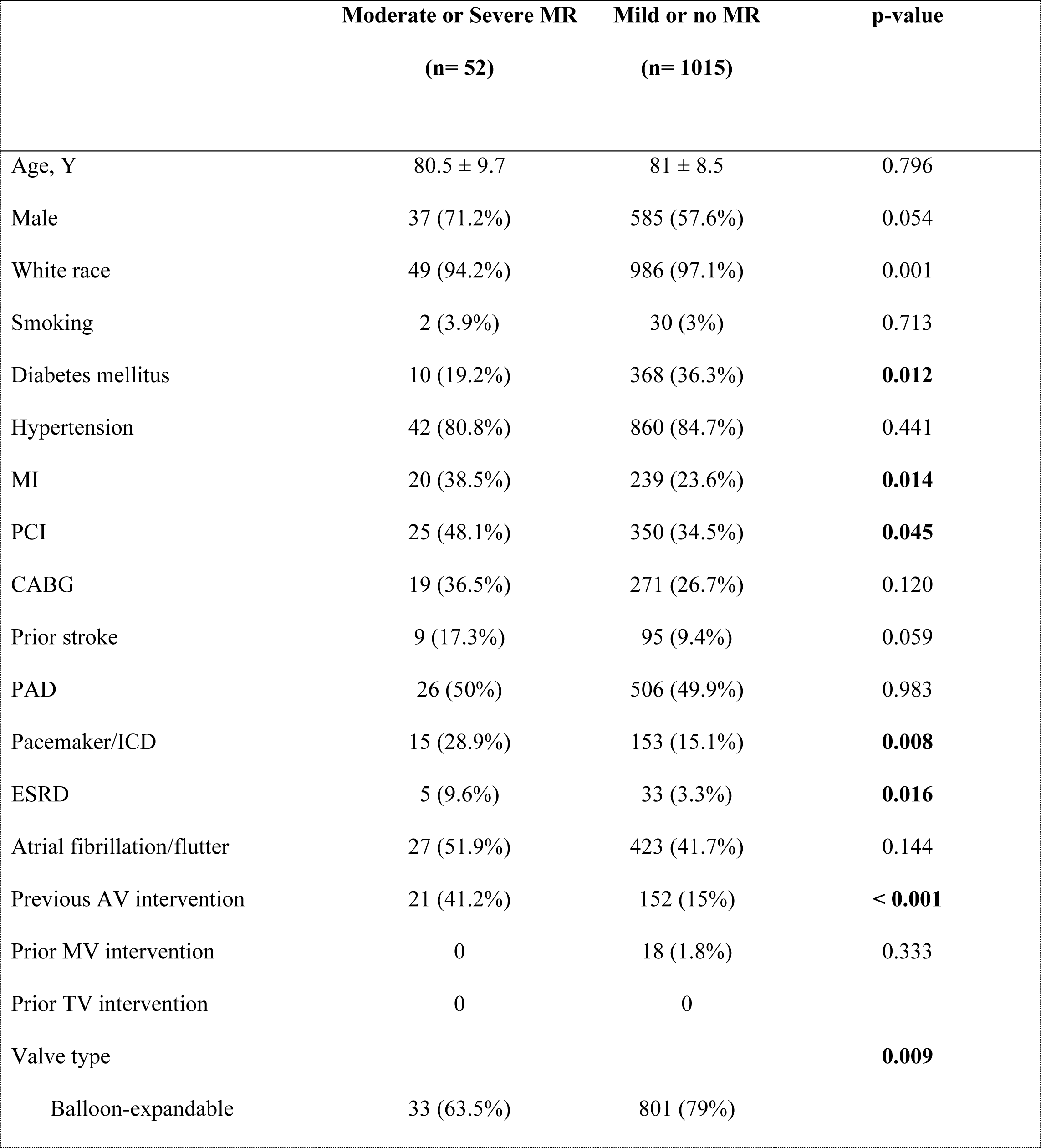
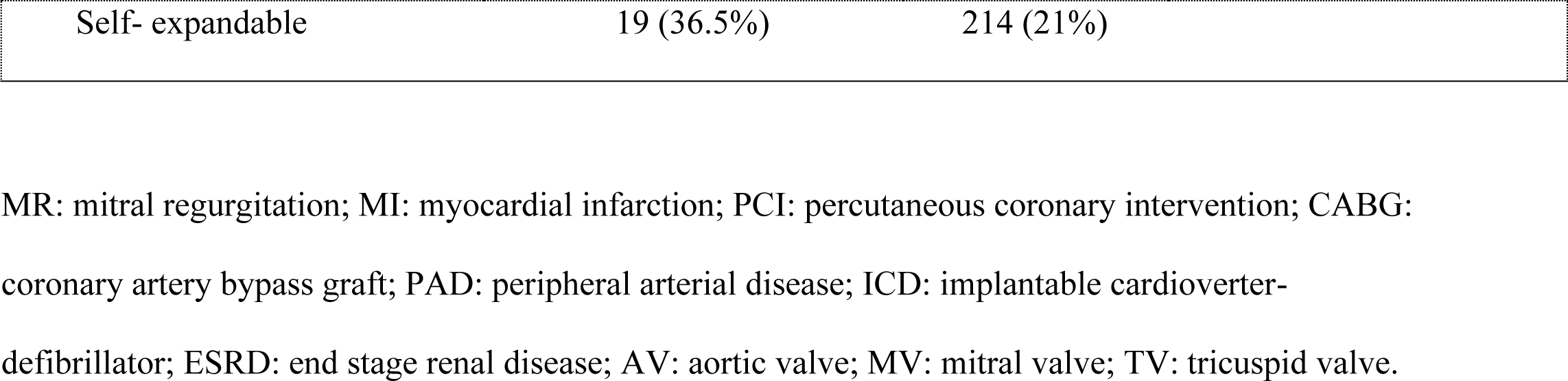
Baseline characteristics of the study patients.

|  | Moderate or Severe MR<br>(n= 52) | Mild or no MR<br>(n= 1015) | p-value |
| --- | --- | --- | --- |
| Age, Y | 80.5 ± 9.7 | 81 ± 8.5 | 0.796 |
| Male | 37 (71.2%) | 585 (57.6%) | 0.054 |
| White race | 49 (94.2%) | 986 (97.1%) | 0.001 |
| Smoking | 2 (3.9%) | 30 (3%) | 0.713 |
| Diabetes mellitus | 10 (19.2%) | 368 (36.3%) | **0.012** |
| Hypertension | 42 (80.8%) | 860 (84.7%) | 0.441 |
| MI | 20 (38.5%) | 239 (23.6%) | **0.014** |
| PCI | 25 (48.1%) | 350 (34.5%) | **0.045** |
| CABG | 19 (36.5%) | 271 (26.7%) | 0.120 |
| Prior stroke | 9 (17.3%) | 95 (9.4%) | 0.059 |
| PAD | 26 (50%) | 506 (49.9%) | 0.983 |
| Pacemaker/ICD | 15 (28.9%) | 153 (15.1%) | **0.008** |
| ESRD | 5 (9.6%) | 33 (3.3%) | **0.016** |
| Atrial fibrillation/flutter | 27 (51.9%) | 423 (41.7%) | 0.144 |
| Previous AV intervention | 21 (41.2%) | 152 (15%) | **< 0.001** |
| Prior MV intervention | 0 | 18 (1.8%) | 0.333 |
| Prior TV intervention | 0 | 0 |  |
| Valve type |  |  | **0.009** |
| Balloon-expandable | 33 (63.5%) | 801 (79%) |  |

|  |  |  |
| --- | --- | --- |
| Self- expandable | 19 (36.5%) | 214 (21%) |

MR: mitral regurgitation; MI: myocardial infarction; PCI: percutaneous coronary intervention; CABG: coronary artery bypass graft; PAD: peripheral arterial disease; ICD: implantable cardioverter-defibrillator; ESRD: end stage renal disease; AV: aortic valve; MV: mitral valve; TV: tricuspid valve.

Patients with moderate/severe MR were more likely to have prior myocardial infarction (38.5% vs. 23.6%, p = 0.014) and end-stage renal disease (9.6% vs. 3.3%, p = 0.016) compared to those with mild/no MR. On the other hand, they had less diabetes mellitus (19.2% vs. 36.3%, p = 0.012) and were less likely to receive balloon-expandable aortic valve prosthesis (63.5% vs. 79%, p=0.009). Outcomes of the study are summarized in [**Table 2].** There was no significant difference between the moderate/severe MR and mild/no MR groups in procedure success rate, and in-hospital all-cause mortality. At 1- year follow up, there were 8 patients died in the moderate/severe MR group, and 143 patients died in mild/no MR group with no statistically significant difference (15.4% vs. 14.1%, p=794) **[Figure 1]**. In multivariate analysis, moderate/severe MR was not associated with higher all-cause mortality at 1-year follow up (OR 0.88, 95% CI [0.39, 1.99], p=0.761) **[Table 3]**.

**Figure 1.**
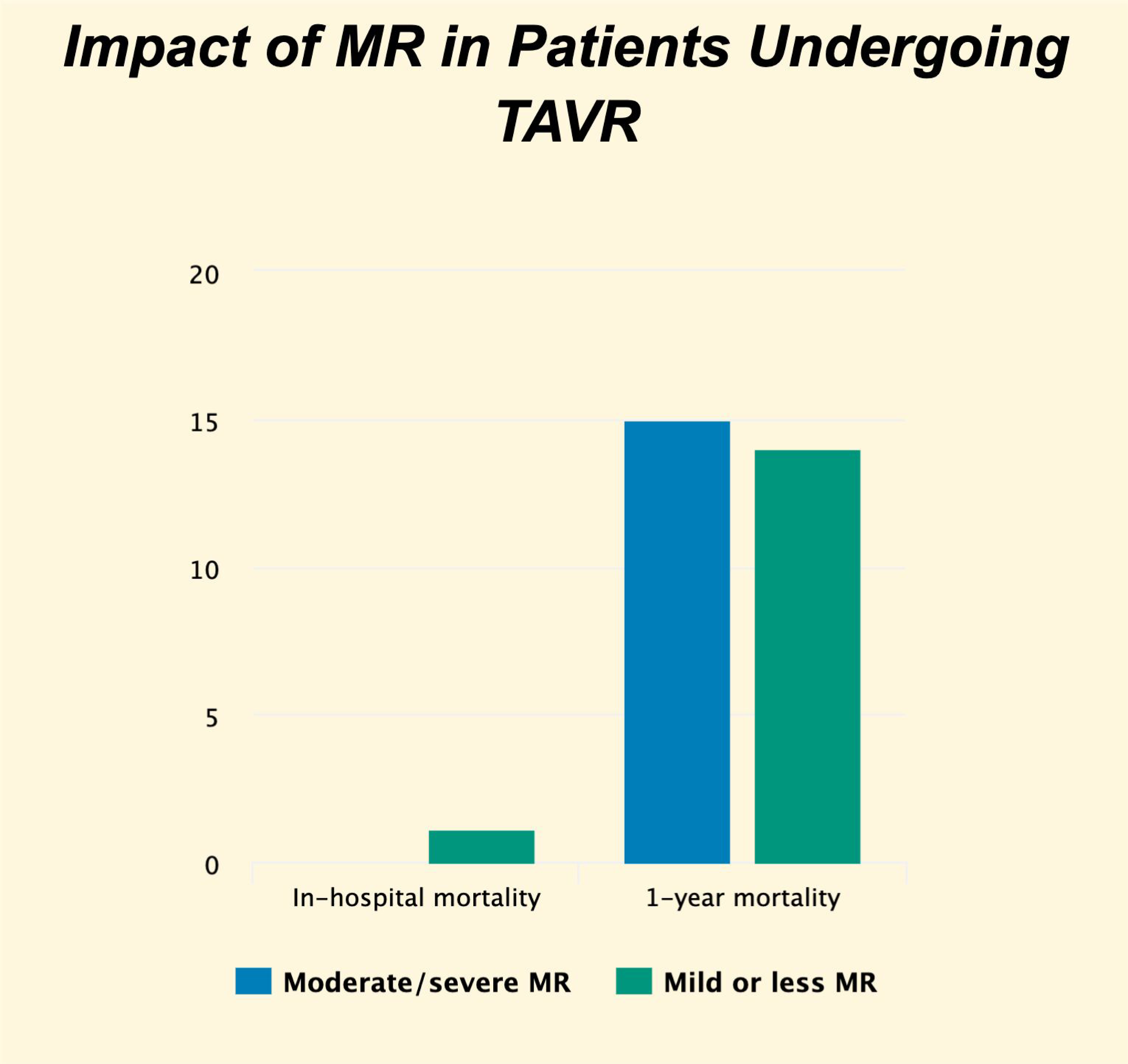
Impact of moderate/severe mitral regurgitation in all-cause mortality during hospitalization and at 1-year follow up post TAVI

**Table 2.** Outcomes of moderate/severe valvular regurgitation.

|  | **Moderate or Severe MR** | **Mild or no MR** | **P value** |
| --- | --- | --- | --- |
|  | **(n= 52)** | **(n= 1015)** |  |
| TAVI procedure Success | 52 (100%) | 993 (97.8%) | 0.283 |
| In-hospital all-cause mortality | 0 | 11 (1.1%) | 0.450 |
| One-year all-cause mortality | 8 (15.4%) | 143 (14.1%) | 0.794 |
|  | **Moderate or Severe TR** | **Mild or no TR** | **P value** |
|  | **(n= 86)** | **(n= 985)** |  |
| TAVI procedure Success | 85 (98.8%) | 964 (97.9%) | 0.543 |
| In-hospital all-cause mortality | 0 | 11 (1.1%) | 0.325 |
| One-year all-cause mortality | 23 (26.7%) | 130 (13.2%) | **0.001** |

MR: mitral regurgitation; TR: tricuspid regurgitation; TAVI: transcatheter aortic valve implantation.

**Table 3.** Multivariate analysis in patients underwent TAVI.

|  | **Odds ratio** | **Standard error** | **P value** | **95% Confidence interval** |
| --- | --- | --- | --- | --- |
| **Moderate/severe TR** | 1.944607 | 0.5675503 | **0.023** | **1.097492 - 3.44558** |
| **Moderate/severe MR** | 0.880866 | 0.3675831 | 0.761 | 0.3887815 - 1.995787 |
| **Gender** | 1.030002 | 0.2076312 | 0.883 | 0.6938252 - 1.529065 |
| **Age** | 1.016118 | 0.0117927 | 0.168 | 0.9932659 - 1.039496 |
| **Prior MV intervention** | 1.406484 | 0.850043 | 0.573 | 0.4302195 - 4.598109 |
| **Hypertension** | 0.8124038 | 0.2149982 | 0.432 | 0.4836221 - 1.364702 |
| **Diabetes mellitus** | 1.053106 | 0.208657 | 0.794 | 0.714199 - 1.552833 |
| **ESRD** | 4.505258 | 1.675088 | **<0.001** | **2.173885 - 9.336898** |
| **Prior MI** | 1.031806 | 0.2429494 | 0.894 | 0.6503901 - 1.6369 |
| **Atrial fibrillation/flutter** | 2.062502 | 0.413216 | **<0.001** | **1.392703 - 3.054429** |
| **Prior CABG** | 1.019374 | 0.2342097 | 0.933 | 0.6497747 - 1.599205 |
| **Pacemaker/ICD** | 0.8736883 | 0.2215863 | 0.594 | 0.5314634 - 1.436282 |
| **TAVI procedure success** | 0.1817218 | 0.0847071 | **<0.001** | **0.0728839 - 0.4530882** |

TAVI: transcatheter aortic valve implantation; TR: tricuspid regurgitations; MR: mitral regurgitation;
MV: mitral valve; ESRD: end stage renal disease; MI: myocardial infarction; CABG: coronary artery
bypass graft; ICD: implantable cardioverter-defibrillator.

### Comparing moderate/severe TR vs. mild/no TR

The baseline demographics and patient characteristics were summarized in [**Table 4].** The moderate/severe TR group included 86 patients (8%), and the mild/no TR group included 985 patients (92%), with a total of 1,071 patients. The mild/no TR group had a greater proportion of males (59.39% vs. 45.35%, p = 0.011) compared to moderate/severe TR group. Patients with moderate/severe TR had lower incidence of diabetes mellitus (19.8% vs. 36.8%, p=0.002), and percutaneous coronary interventions (24.4% vs. 35.9%, p=0.032), but higher incidence of atrial fibrillation (80.2% vs. 38.8%, p<0.001), and prior mitral valve intervention (5.8% vs. 1.4%, p=0.03). There was no difference in procedural success rate or in-hospital all-cause mortality between both groups **[Table 2]**. At 1-year follow up, all-cause mortality was significantly higher in moderate/severe TR group compared to mild/no TR group (26.7% vs. 13.2%, p=0.001) **[Table 2] [Figure 2]**. In multivariate analysis, moderate/severe TR was associated with higher all-cause mortality at 1-year follow up (OR 1.94, 95% CI [01.09, 3.44], p=0.0.023) **[Table 3]**.

**Figure 2.**
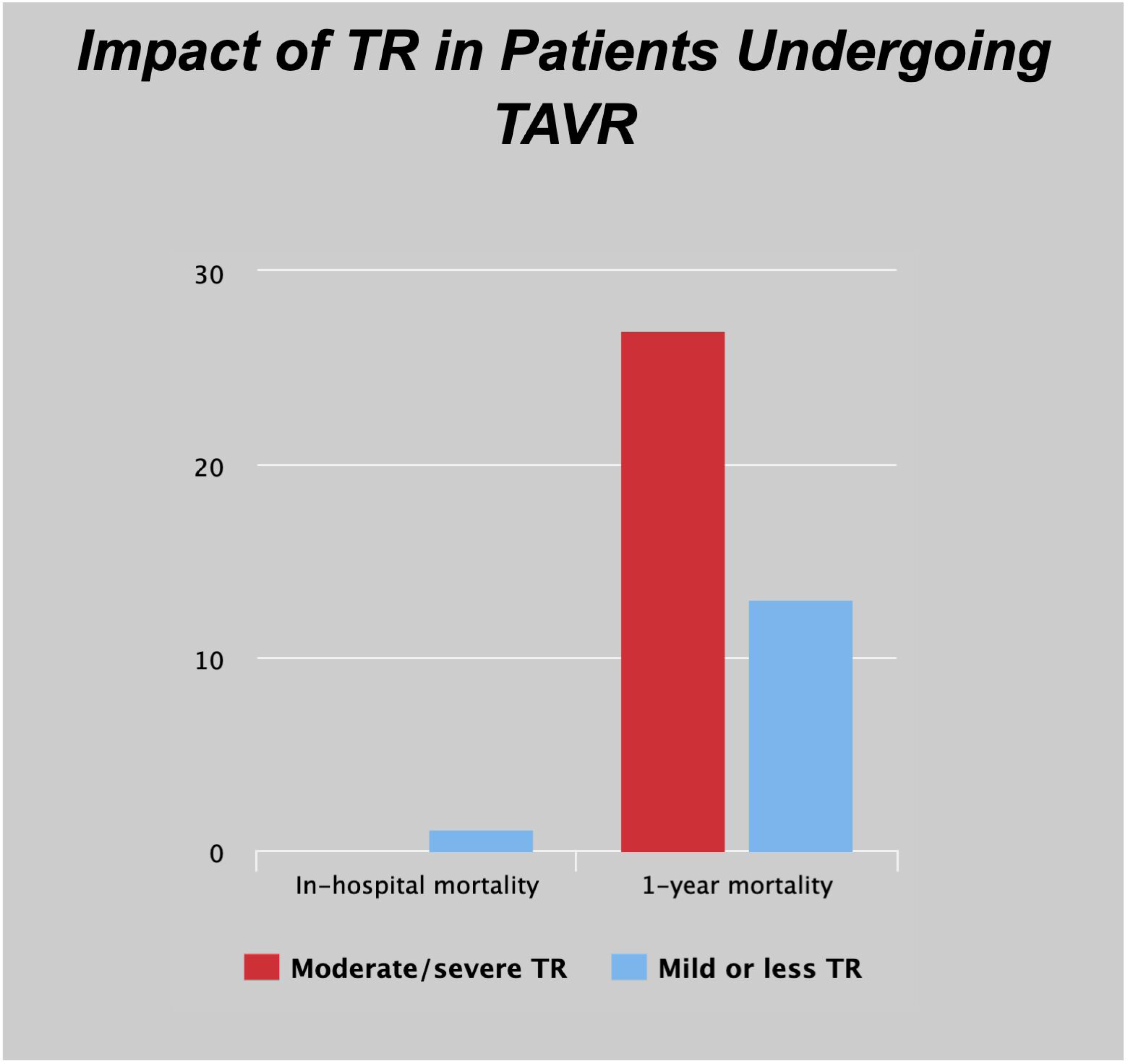
Impact of moderate/severe tricuspid regurgitation in all-cause mortality during hospitalization and at 1-year follow up post TAVI

**Table 4.**
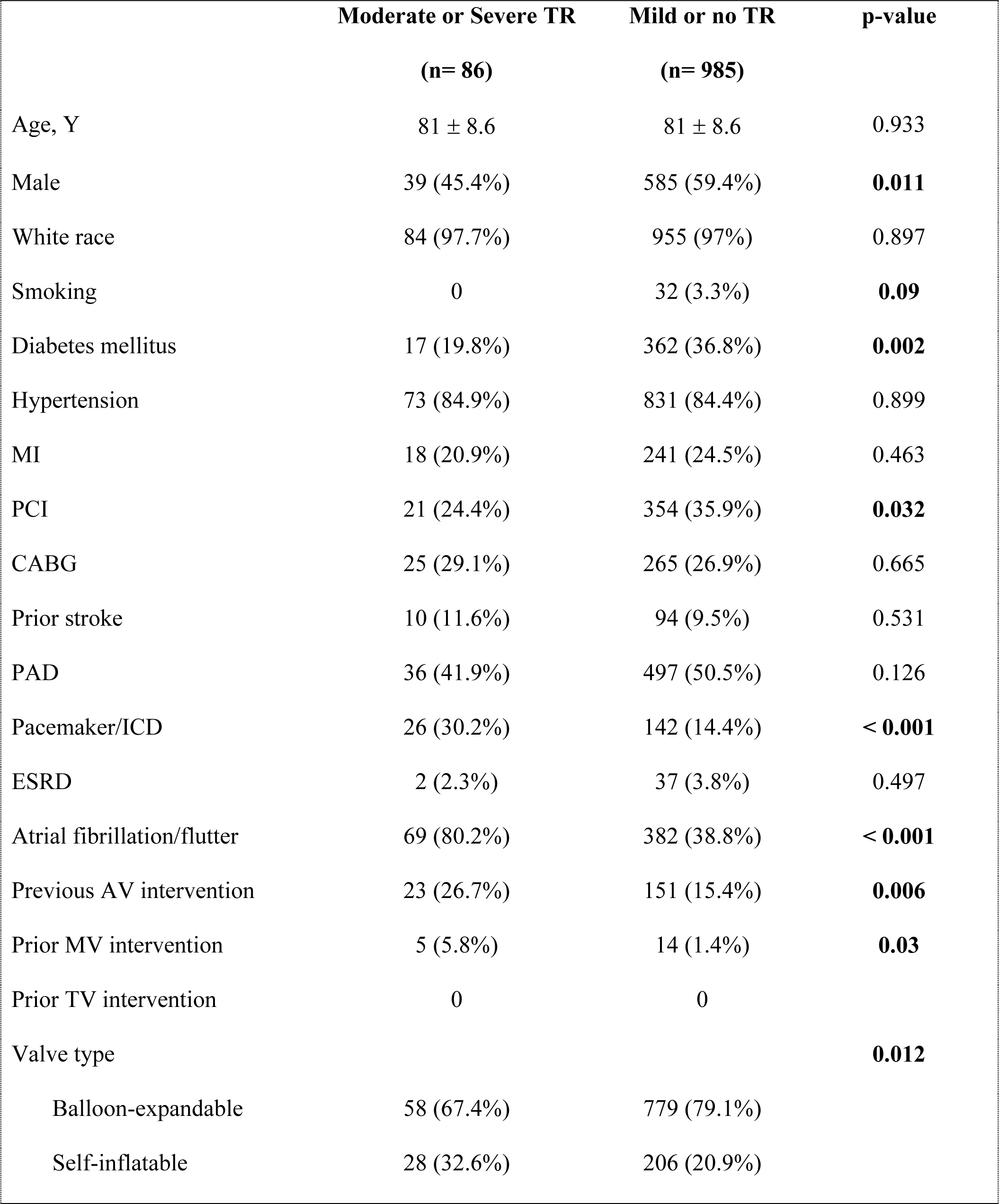
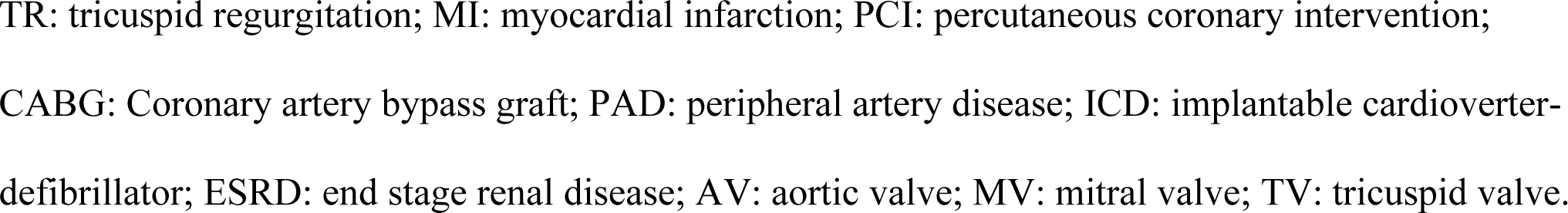
Baseline characteristics of the study patients.

|  | Moderate or Severe TR<br>(n= 86) | Mild or no TR<br>(n= 985) | p-value |
| --- | --- | --- | --- |
| Age, Y | 81 ± 8.6 | 81 ± 8.6 | 0.933 |
| Male | 39 (45.4%) | 585 (59.4%) | **0.011** |
| White race | 84 (97.7%) | 955 (97%) | 0.897 |
| Smoking | 0 | 32 (3.3%) | **0.09** |
| Diabetes mellitus | 17 (19.8%) | 362 (36.8%) | **0.002** |
| Hypertension | 73 (84.9%) | 831 (84.4%) | 0.899 |
| MI | 18 (20.9%) | 241 (24.5%) | 0.463 |
| PCI | 21 (24.4%) | 354 (35.9%) | **0.032** |
| CABG | 25 (29.1%) | 265 (26.9%) | 0.665 |
| Prior stroke | 10 (11.6%) | 94 (9.5%) | 0.531 |
| PAD | 36 (41.9%) | 497 (50.5%) | 0.126 |
| Pacemaker/ICD | 26 (30.2%) | 142 (14.4%) | **< 0.001** |
| ESRD | 2 (2.3%) | 37 (3.8%) | 0.497 |
| Atrial fibrillation/flutter | 69 (80.2%) | 382 (38.8%) | **< 0.001** |
| Previous AV intervention | 23 (26.7%) | 151 (15.4%) | **0.006** |
| Prior MV intervention | 5 (5.8%) | 14 (1.4%) | **0.03** |
| Prior TV intervention | 0 | 0 |  |
| Valve type |  |  | **0.012** |
| Balloon-expandable | 58 (67.4%) | 779 (79.1%) |  |
| Self-inflatable | 28 (32.6%) | 206 (20.9%) |  |

TR: tricuspid regurgitation; MI: myocardial infarction; PCI: percutaneous coronary intervention; CABG: Coronary artery bypass graft; PAD: peripheral artery disease; ICD: implantable cardioverter- defibrillator; ESRD: end stage renal disease; AV: aortic valve; MV: mitral valve; TV: tricuspid valve.

## Discussion

In this retrospective observational study, we investigated the impact of concomitant moderate/severe MR and TR in patients who underwent TAVI. In our cohort, 4.9% of the patients had concomitant moderate/severe MR and 8% had concomitant moderate/severe TR. The key findings of our analysis are summarized in the following: (1) Moderate/severe TR was associated with higher all-cause mortality at 1-year follow up; (2) Moderate to severe MR did not affect mortality at follow up; (3) Neither significant MR nor TR affected the TAVI procedure success or in-hospital death; (4) In multivariate analysis, moderate/severe TR was associated with 2-fold higher mortality at 1-year follow up.

Aortic stenosis is notably prevalent in the elderly population, as evidenced by a prospective study highlighting a notable rise in its incidence from 0.2% among individuals in their 50s to a significant 9.8% among those in their 80s. (13). The etiology of concomitant valvular dysfunction in patients with severe AS is frequent and often multifactorial. Significant aortic stenosis obstructs blood flow and reduces forward flow causing left ventricular pressure build-up, and subsequently elevated filling pressures in left ventricle and left atrium which in-turn causes pulmonary hypertension, right sided heart failure and significant functional MR and TR. In addition, cardiac remodeling induced by severe AS leads to functional MR (14). Smoking, hypertension, diabetes, peripheral arterial diseases, and ESRD, significantly impact the progression of AS, MR, and TR. AS advances rapidly in individuals with ESRD due to increased metabolic and hemodynamic stress on the aortic valve, resulting in an accelerated disease course and a poorer prognosis even with mild-to-moderate AS in ESRD (15).

Current guidelines incorporate multiple factors in risk stratifying patients with severe AS such as: echocardiographic parameters, presence of AS-related symptoms, and multiple other comorbidities (16). However, concomitant other valvular dysfunction was not one of the factors. Genereux et al. proposed AS staging based on the extent of cardiac damage, giving moderate/severe MR stage 2 and moderate/severe TR stage 3 (17). The higher the stage, the worse the prognosis after TAVI. In our analysis, baseline moderate to severe TR was associated with 2-folds higher mortality at 1-year follow up post TAVI procedure. In other analysis, progression of mild or less to moderate or higher TR post TAVI was also associated with higher mortality at 2-year follow-up, likely related to worsening RV function and pulmonary hypertension (18). In addition, previous meta-analyses had linked significant TR, RV dysfunction, and pulmonary hypertension with higher mortality and worse prognosis post TAVI (19,20).

Data about prognostic significance of MR post TAVI is controversial. In our analysis, baseline significant MR was not associated with higher mortality either in-hospital or at 1-year follow up. A finding that was confirmed in the multivariate analysis. TAVI may improve LV stroke volume and unload the LV, as result it may improve MR after the procedure. In one study, 59% of patients with significant MR regressed after TAVI (21). In the SWEDEHEART registry, the persistence or worsening of significant MR was associated with higher mortality post TAVI. However, MR improvement post TAVI offset this effect (22). In Baz et al. persistent significant MR at 6-months post TAVI was an independent predictor of mortality at 2-years (23). However, in our analysis we did not include transthoracic echo data post TAVI and at follow up. As a result, a careful transthoracic echo assessment pre and post TAVI is warranted as it may affect patients’ prognosis and risk stratification.

## Limitations

Our study has few limitations, including those inherent to retrospective observational study design. In addition, dependence on the Mayo enterprise database adds further restrictions. There were multiple differences in the demographics and baseline patients’ characteristics between the two arms of our study, which may affect the outcomes, but we conducted a multivariate analysis to correct for the differences and demonstrate the independent predictors of mortality at 1-year follow up post TAVI.

## Conclusion

Our study reveals that moderate or severe TR is associated with a significant increase in all-cause mortality at one year after TAVI. However, at the same period, moderate or severe MR did not demonstrate a significant association with mortality. These findings emphasize the importance of thorough assessment and consideration of multivalvular disease in TAVI patients to optimize long-term outcomes. Future research should address these limitations to enhance treatment strategies and refine clinical decision-making.

## Data Availability

Data is available

## References

1 Steeds RP, Potter A, Mangat N, et al. Community-based aortic stenosis detection: clinical and echocardiographic screening during influenza vaccination. Open Heart. 2021;8(1):e001640. doi:10.1136/openhrt-2021-001640

2 Haensig M, Holzhey DM, Borger MA, et al. Improved mitral valve performance after transapical aortic valve implantation. Ann Thorac Surg. 2014;97(4):1247–1254. doi:10.1016/j.athoracsur.2013.11.025

3 Perl L, Vaturi M, Assali A, et al. The Impact of Transcatheter Aortic Valve Implantation on Mitral Regurgitation Regression in High-Risk Patients with Aortic Stenosis. J Heart Valve Dis. 2015;24(4):439–444.

4 Zilberszac R, Gleiss A, Binder T, et al. Prognostic relevance of mitral and tricuspid regurgitation in patients with severe aortic stenosis. Eur Heart J Cardiovasc Imaging. 2018;19(9):985–992. doi:10.1093/ehjci/jey027

5 Barbanti M, Binder RK, Dvir D, et al. Prevalence and impact of preoperative moderate/severe tricuspid regurgitation on patients undergoing transcatheter aortic valve replacement. Catheter Cardiovasc Interv. 2015;85(4):677–684. doi:10.1002/ccd.25512

6 Hutter A, Bleiziffer S, Richter V, et al. Transcatheter aortic valve implantation in patients with concomitant mitral and tricuspid regurgitation. Ann Thorac Surg. 2013;95(1):77–84. doi:10.1016/j.athoracsur.2012.08.030

7 Amat-Santos IJ, Castrodeza J, Nombela-Franco L, et al. Tricuspid but not Mitral Regurgitation Determines Mortality After TAVI in Patients With Nonsevere Mitral Regurgitation. Rev Esp Cardiol (Engl Ed). 2018;71(5):357–364. doi:10.1016/j.rec.2017.08.019

8 Alaour B, Nakase M, Pilgrim T. Combined Significant Aortic Stenosis and Mitral Regurgitation: Challenges in Timing and Type of Intervention. Can J Cardiol. Published online November 4, 2023. doi:10.1016/j.cjca.2023.11.003

9 Matta A, Kanso M, Kibler M, et al. Long-Term Survival Outcomes After Transcatheter Aortic Valve Replacement: A Real-World Experience of a Large Tertiary Center. Am J Cardiol. 2023;207:229–236. doi:10.1016/j.amjcard.2023.09.001

10 Harling L, Saso S, Jarral OA, Kourliouros A, Kidher E, Athanasiou T. Aortic valve replacement for aortic stenosis in patients with concomitant mitral regurgitation: should the mitral valve be dealt with?. Eur J Cardiothorac Surg. 2011;40(5):1087–1096. doi:10.1016/j.ejcts.2011.03.036

11 Leon MB, Smith CR, Mack M, et al. Transcatheter aortic-valve implantation for aortic stenosis in patients who cannot undergo surgery. N Engl J Med. 2010;363(17):1597–1607. doi:10.1056/NEJMoa1008232

12 Kowalówka AR, Onyszczuk M, Wańha W, Deja MA. Do we have to operate on moderate functional mitral regurgitation during aortic valve replacement for aortic stenosis?. Interact Cardiovasc Thorac Surg. 2016;23(5):806–809. doi:10.1093/icvts/ivw212

13 Matta A, Levai L, Roncalli J, et al. Comparison of in-hospital outcomes and long-term survival for valve-in-valve transcatheter aortic valve replacement versus the benchmark native valve transcatheter aortic valve replacement procedure. Front Cardiovasc Med. 2023;10:1113012. Published 2023 Feb 9. doi:10.3389/fcvm.2023.1113012

14 Park SM, Park SW, Casaclang-Verzosa G, et al. Diastolic dysfunction and left atrial enlargement as contributing factors to functional mitral regurgitation in dilated cardiomyopathy: data from the Acorn trial. Am Heart J. 2009;157(4):762.e3- 762.e762010. doi:10.1016/j.ahj.2008.12.018

15 Kim D, Shim CY, Hong GR, et al. Effect of End-Stage Renal Disease on Rate of Progression of Aortic Stenosis. Am J Cardiol. 2016;117(12):1972–1977. doi:10.1016/j.amjcard.2016.03.048

16 Writing Committee Members, et al. “2020 ACC/AHA guideline for the management of patients with valvular heart disease: a report of the American College of Cardiology/American Heart Association Joint Committee on Clinical Practice Guidelines.” journal of the American College of Cardiology 77.4 (2021): e25-e197.

17 Généreux P, Pibarot P, Redfors B, Mack MJ, Makkar RR, Jaber WA, Svensson LG, Kapadia S, Tuzcu EM, Thourani VH, Babaliaros V. Staging classification of aortic stenosis based on the extent of cardiac damage. European heart journal. 2017 Dec 1;38(45):3351–8.

18 Muraishi M, Tabata M, Shibayama K, Ito J, Shigetomi K, Obunai K, Watanabe H, Yamamoto M, Watanabe Y, Naganuma T, Shirai S. Late Progression of Tricuspid Regurgitation After Transcatheter Aortic Valve Replacement. Journal of the Society for Cardiovascular Angiography & Interventions. 2022 May 1;1(3):100043.

19 Kokkinidis DG, Papanastasiou CA, Jonnalagadda AK, Oikonomou EK, Theochari CA, Palaiodimos L, Karvounis HI, Armstrong EJ, Faillace RT, Giannakoulas G. The predictive value of baseline pulmonary hypertension in early and long term cardiac and all-cause mortality after transcatheter aortic valve implantation for patients with severe aortic valve stenosis: A systematic review and meta-analysis. Cardiovascular Revascularization Medicine. 2018 Oct 1;19(7):859–67.

20 Takagi H, Hari Y, Kawai N, Ando T; ALICE (All-Literature Investigation of Cardiovascular Evidence) Group. Impact of concurrent tricuspid regurgitation on mortality after transcatheter aortic-valve implantation. Catheter Cardiovasc Interv. 2019;93(5):946–953. doi:10.1002/ccd.27948

21 Winter MP, Bartko PE, Krickl A, Gatterer C, Donà C, Nitsche C, Koschutnik M, Spinka G, Siller-Matula JM, Lang IM, Mascherbauer J. Adaptive development of concomitant secondary mitral and tricuspid regurgitation after transcatheter aortic valve replacement. European Heart Journal-Cardiovascular Imaging. 2021 Sep 1;22(9):1045–53.

22 Feldt K, De Palma R, Bjursten H, Petursson P, Nielsen NE, Kellerth T, Jönsson A, Nilsson J, Rück A, Settergren M. Change in mitral regurgitation severity impacts survival after transcatheter aortic valve replacement. International Journal of Cardiology. 2019 Nov 1;294:32–6.

23 Bäz L, Möbius-Winkler S, Diab M, Kräplin T, Westphal JG, Ibrahim K, Schulze PC, Franz M. Prognostic relevance of mitral and tricuspid regurgitation after transcatheter aortic valve implantation: Impact of follow-up time point for decision-making. Frontiers in Cardiovascular Medicine. 2023 Feb 16;10:990373.

